# Time-to-event comparative effectiveness of NOACs vs VKAs in newly diagnosed non-valvular atrial fibrillation patients

**DOI:** 10.1101/2021.08.06.21261092

**Authors:** Blanca Gallego, Jie Zhu

## Abstract

**Objective:** To investigate the difference in the time-to-event probabilities of ischaemic events, major bleeding and death of NOAC vs VKAs in newly diagnosed non-valvular atrial fibrillation patients.

**Design:** Retrospective observational cohort study.

**Setting:** UK’s Clinical Practice Research Data linked to the Hospital Episode Statistics inpatient and outpatient data, mortality data and the Patient Level Index of Multiple Deprivation.

**Participants:** Patients over 18 years of age, with an initial diagnosis of atrial fibrillation between 1st-Mar-2011 and 31-July-2017, without a record for a valve condition, prosthesis or procedure previous to initial diagnosis, and without a record of oral anticoagulant treatment in the previous year.

**Intervention:** Oral anticoagulant treatment with either vitamin K antagonists (VKAs) or the newer target-specific oral anticoagulants (NOACs).

**Main Outcome Measures:** Ischaemic event, major bleeding event and death from 15 days from initial prescription up to two years follow-up.

**Statistical Analysis:** Treatment effect was defined as the difference in time-to-event probability between NOAC and VKA treatment groups. Treatment and outcomes were modelled using an ensemble of parametric and non-parametric models, and the average and conditional average treatment effects were estimated using one-step Targeted Maximum Likelihood Estimation (TMLE). Heterogeneity of treatment effect was examined using variable importance methods in Bayesian Additive Regression Trees (BART).

**Results:** The average treatment effect of NOAC vs VKA was consistently close to zero across all times, with a temporal average of 0.00[95%0.00, 0.00] for ischaemic event, 0.00%[95% *−*0.01, 0.01] for major bleeding and 0.00[95% *−*0.01, 0.01] for death. Only history of major bleeding was found to influence the distribution of treatment effect for major bleeding, but its impact on the associated conditional average treatment effect was not significant.

**Conclusions:** This study found no statistically significant difference between NOAC and VKA users up to two years of medication use for the prevention of ischaemic events, major bleeding or death.

## 1 Introduction

Atrial fibrillation (AF) is the most common cardiac arrhythmia and can lead to blood clots forming in the heart. Oral anticoagulant treatment with either vitamin K antagonists (VKAs) or the newer target-specific oral anticoagulants (NOACs), also refer to as direct oral anticoagulants (DOACs), are commonly used in these patients to prevent stroke and other complications (NICE Last Updated: August 2014). The use of oral anticoagulants needs to be balanced against the risk of bleeding. The use of vitamin K antagonists is further limited by a narrow therapeutic index requiring regular monitoring as well as by higher uncertainty in their pharmacokinetic and pharmacodynamic properties.

In the past years, there have been many experimental and observational studies investigating the average comparative efficacy and safety of these anticoagulants. A meta-analysis of four large, phase III clinical trials of patients with non-valvular AF who were randomised to receive NOACs (dabigatran, rivaroxaban, apixaban and edoxaban) or Warfarin (the most commonly used VKA) [19] concluded that NOACs had a favourable risk–benefit profile. They were found to be at least as effective as Warfarin at preventing stroke with the benefit of lower mortality and a reduction against haemorrhagic stroke and intracranial bleeding, albeit with a higher incidence of gastrointestinal bleeding. A more recent meta-analysis of 23 RCTs (13 comparing NOACs with Warfarin) [11] arrived at similar findings although inspection of the 95% confidence intervals indicate that, in most cases, the reductions of risk associated with some NOACs were not statistically significant, with the exception of intracranial bleeding for which all NOACs offered statistically significant superior protection. Another systematic review and meta-analyses, this time of 28 observational studies, [**?**] compared the safety and efficacy of three NOACs (dabigatran, rivaroxaban and apixaban) vs VKAs. This meta-analysis confirmed that NOACs in general have similar rates of ischaemic stroke or systemic embolism and reduced intracranial haemorrhage as compared to VKAs. Apixaban and dabigatran were associated with lower mortality and dabigatran and rivaroxaban with higher risk of gastrointestinal bleeding.

Less evidence exists regarding the optimal choice of therapy based on individual patient characteristics. Secondary analysis of the large clinical trials comparing NOACs with VKAs are not conclusive. A subgroup analysis of the large clinical trials comparing NOACs with warfarin found no heterogeneity in important subgroups, with the exception of time in therapeutic range <66%, which was associated with a greater relative reduction in major bleeding with NOACs vs warfarin [19]. However, a pooled analysis of gender differences in residual stroke and major bleeding in patients treated with warfarin or NOAC from clinical trial studies found women (but not men) treated with NOAC vs warfarin had a reduction in major bleeding event [14]. No further gender differences were found in an indirect comparison amongst the various NOACs [12].

All previous observational studies were characterised by providing a treatment effect defined as a Hazard Ratio using Cox models. In this observational cohort study we extend current methodology by defining treatment effect as the difference in time-to-event (survival) probability between NOAC and VKA treatment groups and not as a Relative Risk measure or a Hazard Ratio under Cox proportionality assumptions. This absolute measure of risk is more appropriate for clinical decision support since the presence of heterogeneity on relative measures does not imply clinically meaningful heterogeneity [8]. Furthermore, by modelling both event and censor data, potential time-dependent biases associated with loss to follow-up were taken into account.

## 2 Methods

### 2.1 Data sources

Study data was obtained from the UK’s Clinical Practice Research Datalink (CPRD) and included the primary care database (GOLD) linked to the Hospital Episode Statistics (HES) Admitted Patient Care, the HES Outpatient Data, the Office for National Statistics (ONS) Mortality data and the Patient Level Index of Multiple Deprivation.

The CPRD GOLD database contains routinely collected primary care records from over 20 million patient lives, with over 5 million currently registered and active patients, representative of the UK population with respect to age, gender and ethnicity. The quality of data in CPRD is subject to rigorous checks and is monitored regularly. The HES data contains information of all admissions to National Health Service hospitals in England including detailed admission and outpatient administrative data. The ONS mortality data contains the date and cause of death for the population of England and Wales. Over 50% of patients found in the CPRD GOLD are linked to HES and ONS. Detailed information on the content, quality and access to this data can be found at CPRD data resource profile [22].

### 2.2 Study design

#### Study cohort

An initial cohort was selected as all eligible patients with a diagnosis of AF (either in CPRD or HES datasets) between 1st-Mar-2011 and 31-July-2017. Patients were eligible if they were: 18 years or over at time of first diagnosis, met CPRD’s research quality indicators, had been registered with CPRD for at least one year before their initial diagnosis, and had their records linked to HES data. Patients with a history of AF or with a record for a valve condition, prosthesis or procedure previous to initial diagnosis were excluded from the study. A complete list of inclusion and exclusion criteria codes can be found in Supplementary Table 1.

**Table 1:** Baseline characteristics

| Variable | VKA (n=13,159) |  |  |  |  | NOAC (n=7,111) |  |  |  |  | OR [CI95%]* | p-value |
| --- | --- | --- | --- | --- | --- | --- | --- | --- | --- | --- | --- | --- |
|  | n (0) | % (0) | 25% | 50% | 75% | n | % | 25% | 50% | 75% |  |  |
| Age | 13159 | 100 | 68 | 76 | 82 | 7111 | 100 | 69 | 77 | 84 | 1.01 [1.01,1.01] | 0.001 |
| Female | 5775 | 43.9 | 0 | 0 | 1 | 3219 | 45 | 0 | 0 | 1 | 1.06 [1.00,1.12] | 0.059 |
| Deprivation index | 13159 | 100 | 3 | 5 | 7 | - | - | 2 | 4 | 7 | 0.97 [0.96,0.99] | 0.001 |
| Paroxysmal AF | 31 | 0.2 | 0 | 0 | 0 | 204 | 2.9 | 0 | 0 | 0 | 12.51 [8.70,18.62] | 0.001 |
| Persistent/Chronic AF | 27 | 0.2 | 0 | 0 | 0 | 19 | 0.3 | 0 | 0 | 0 | 1.30 [0.71,2.33] | 0.377 |
| First AF at hospital | 5880 | 44.7 | 0 | 0 | 1 | 3335 | 47 | 0 | 0 | 1 | 1.09 [1.03,1.16] | 0.003 |
| Labile INR | 1344 | 10.2 | 0 | 0 | 0 | 55 | 0.8 | 0 | 0 | 0 | 0.17 [0.13,0.20] | 0.001 |
| Charlson index | 13007 | 98.8 | 4 | 5 | 6 | 7030 | 99 | 4 | 5 | 7 | 1.02 [1.01,1.03] | 0.001 |
| Chad2vasc2 score | 13017 | 98.9 | 3 | 4 | 5 | 7056 | 99 | 3 | 4 | 5 | 1.04 [1.02,1.06] | 0.001 |
| Hasbled score | 13050 | 99.2 | 2 | 3 | 4 | 7064 | 99 | 2 | 3 | 4 | 0.90 [0.88,0.92] | 0.001 |
| History of stroke | 1781 | 13.5 | 0 | 0 | 0 | 1219 | 17 | 0 | 0 | 0 | 1.07 [1.05,1.10] | 0.001 |
| History of bleeding | 1095 | 8.3 | 0 | 0 | 0 | 608 | 8.6 | 0 | 0 | 0 | 0.98 [0.93,1.03] | 0.4667 |
| Congestive heart failure | 3196 | 24.3 | 0 | 0 | 0 | 1579 | 22 | 0 | 0 | 0 | 1.00 [0.99,1.01] | 0.769 |
| Myocardial infarction | 1684 | 12.8 | 0 | 0 | 0 | 773 | 11 | 0 | 0 | 0 | 0.97 [0.95,0.99] | 0.009 |
| Vascular disease | 817 | 6.2 | 0 | 0 | 0 | 392 | 5.5 | 0 | 0 | 0 | 0.98 [0.94,1.01] | 0.180 |
| Cerebrovascular disease | 1597 | 12.1 | 0 | 0 | 0 | 1153 | 16 | 0 | 0 | 0 | 1.09 [1.06,1.11] | 0.001 |
| Hypertension | 12127 | 92.2 | 6 | 27 | 51 | 6585 | 93 | 5 | 26 | 49 | 1.00 [1.00,1.00] | 0.082 |
| Diabetes (complicated) | 1885 | 14.3 | 0 | 0 | 0 | 1076 | 15 | 0 | 0 | 0 | 1.02 [1.00,1.04] | 0.106 |
| Diabetes (uncomplicated) | 2290 | 17.4 | 0 | 0 | 0 | 1248 | 18 | 0 | 0 | 0 | 1.00 [1.00,1.00] | 0.415 |
| Chronic Kidney disease | 2382 | 18.1 | 0 | 0 | 0 | 1350 | 19 | 0 | 0 | 0 | 0.99 [0.99,1.00] | 0.056 |
| End stage renal failure | 69 | 0.5 | 0 | 0 | 0 | 12 | 0.2 | 0 | 0 | 0 | 0.99 [0.97,1.00] | 0.085 |
| Cancer (any malignancy) | 2935 | 22.3 | 0 | 0 | 0 | 1611 | 23 | 0 | 0 | 0 | 1.01 [1.00,1.02] | 0.063 |
| Cancer (metastatic) | 109 | 0.8 | 0 | 0 | 0 | 79 | 1.1 | 0 | 0 | 0 | 1.00 [0.99,1.02] | 0.717 |
| Liver disease (mild) | 89 | 0.7 | 0 | 0 | 0 | 73 | 1 | 0 | 0 | 0 | 1.03 [0.92,1.15] | 0.577 |
| Liver disease (severe) | 33 | 0.25 | 0 | 0 | 0 | 16 | 0.2 | 0 | 0 | 0 | 0.96 [0.67,1.31] | 0.786 |
| COPD | 2808 | 21.3 | 0 | 0 | 0 | 1551 | 22 | 0 | 0 | 0 | 1.01 [1.00,1.01] | 0.010 |
| Rheumatological disease | 538 | 4.1 | 0 | 0 | 0 | 287 | 4 | 0 | 0 | 0 | 1.00 [0.97,1.03] | 0.796 |
| Gastro intestinal ulcer | 164 | 1.2 | 0 | 0 | 0 | 88 | 1.2 | 0 | 0 | 0 | 0.97 [0.86,1.07] | 0.536 |
| Hemiplegia | 113 | 0.9 | 0 | 0 | 0 | 105 | 1.5 | 0 | 0 | 0 | 1.22 [1.09,1.38] | 0.001 |
| Dementia | 183 | 1.4 | 0 | 0 | 0 | 276 | 3.9 | 0 | 0 | 0 | 1.27 [1.21,1.35] | 0.001 |
| AIDS | 1 | 0.0 | 0 | 0 | 0 | 0 | 0 | 0 | 0 | 0 | - | - |
| Alcoholism | 640 | 4.9 | 0 | 0 | 0 | 421 | 5.9 | 0 | 0 | 0 | 1.08 [1.02,1.14] | 0.007 |
| Aspirin | 7194 | 54.7 | 0 | 1 | 11 | 3069 | 43 | 0 | 0 | 11 | 1.00 [1.00,1.01] | 0.020 |
| NSAID | 3524 | 26.8 | 0 | 0 | 1 | 1683 | 24 | 0 | 0 | 0 | 1.00 [0.99,1.00] | 0.611 |
| Antiplatelets* | 1764 | 13.4 | 0 | 0 | 0 | 1097 | 15 | 0 | 0 | 0 | 1.01 [1.01,1.02] | 0.000 |
| Betablockers | 8675 | 65.9 | 0 | 1 | 11 | 4794 | 67 | 0 | 1 | 12 | 1.00 [1.00,1.01] | 0.001 |
| Antihypertensives | 10525 | 80.0 | 2 | 20 | 39 | 5582 | 79 | 1 | 18 | 35 | 1.00 [1.00,1.00] | 0.260 |

Table 1 continued from previous page VKA (n=13,159) NOAC (n=7,111)
| Variable | Distribution |  |  |  |  | Distribution |  |  |  |  | OR [CI95%]* | p-value |
| --- | --- | --- | --- | --- | --- | --- | --- | --- | --- | --- | --- | --- |
|  | n (0) | % (0) | 25% | 50% | 75% | n | % | 25% | 50% | 75% |  |  |
| Antiarrhythmics | 994 | 7.6 | 0 | 0 | 0 | 411 | 5.8 | 0 | 0 | 0 | 1.02 [1.00,1.03] | 0.018 |
| Antidiabetics | 1710 | 13.0 | 0 | 0 | 0 | 929 | 13 | 0 | 0 | 0 | 1.00 [1.00,1.00] | 0.240 |
| Statins | 7118 | 54.1 | 0 | 2 | 14 | 3969 | 56 | 0 | 2 | 14 | 1.01 [1.00,1.01] | 0.001 |
| SSR uptake Inhibitors | 1327 | 10.1 | 0 | 0 | 0 | 760 | 11 | 0 | 0 | 0 | 1.01 [1.00,1.01] | 0.001 |
| Anticonvulsants | 164 | 1.2 | 0 | 0 | 0 | 66 | 0.9 | 0 | 0 | 0 | 1.00 [0.99,1.00] | 0.325 |
| Proton Pump Inhibitors | 6231 | 47.4 | 0 | 0 | 8 | 3527 | 50 | 0 | 0 | 11 | 1.01 [1.01,1.01] | 0.001 |
| Parenteral Anticoag. | 426 | 3.2 | 0 | 0 | 0 | 157 | 2.2 | 0 | 0 | 0 | 0.94 [0.89,1.00] | 0.041 |
| Fibrinolytic Drugs | 0 | 0.0 | 0 | 0 | 0 | 0 | 0 | 0 | 0 | 0 | - | - |
| Rifampicin | 11 | 0.1 | 0 | 0 | 0 | 5 | 0.1 | 0 | 0 | 0 | 1.06 [0.96,1.33] | 0.378 |
| CP450 Inhibitors | 244 | 1.9 | 0 | 0 | 0 | 128 | 1.8 | 0 | 0 | 0 | 1.05 [0.98,1.12] | 0.175 |
| Pacemaker proc. | 240 | 1.8 | 0 | 0 | 0 | 127 | 1.8 | 0 | 0 | 0 | 1.03 [0.88,1.20] | 0.699 |
| Ablation** | 0 | 0.0 | 0 | 0 | 0 | 0 | 0 | 0 | 0 | 0 | - | - |
| Appendage occlusion** | 0 | 0.0 | 0 | 0 | 0 | 0 | 0 | 0 | 0 | 0 | - | - |
| Cardioversion** | 0 | 0.0 | 0 | 0 | 0 | 0 | 0 | 0 | 0 | 0 | - | - |
| Days from AF diagnosis | 11636 | 88.4 | 12 | 33 | 86 | 6101 | 86 | 9 | 29 | 127.5 | 1.00 [1.00,1.00] | 0.001 |

n and % refer to counts and percentage fraction when variable>0; 25%,50% and 75% are the quantiles of the distribution of the variable.
Lighter grey refers to statistically significant difference with higher prevalence in patients treated with NOACs.
Darker grey refers to statistically significant difference with higher prevalence in patients treated with VKAs.
\*Univariate OR; \*\*Zero at baseline by study design.

#### Interventions

Anticoagulant exposure was determined from prescription records in CPRD. Drug substances associated with VKA exposure included Warfarin sodium (accounting for the large majority of VKA prescriptions), Acenocoumarol, and Phenindione. NOAC exposure included Rivaroxaban, Dabigatran, Edoxaban and Apixaban. A complete list of anticoagulant codes can be found in Supplementary Table 2.

#### Outcomes

We measured four outcomes: (1) death; (2) ischaemic event defined as ischaemic stroke, cerebral arterial occlusion, transient ischaemic attack, other generalised ischaemic cerebrovascular disease, or arterial embolism and thrombosis; and (3) major bleeding defined as haemorrhagic stroke, non-traumatic intracranial haemorrhage, Haemorrhagic disorders, acute bleeding diagnosis and non-traumatic bleeding events during hospitalisation; and (4) a combined outcome of any event (1)-(3). Unidentified stroke was considered to be ischaemic since this is by far the most common type of stroke, with the exception of stroke in the puerperium which is most often haemorrhagic. A complete list of codes related to ischaemic and bleeding events can be found in Supplementary Table 3.

#### Risk Scores and Other Covariates

Patient comorbidities were ascertained by counting the presence of selected diagnosis codes and selected medication codes in two years of medical history from baseline date. The selected diagnoses included common comorbidities such as hypertension, diabetes, congestive heart failure, cancer, chronic pulmonary disease or dementia, and previous stroke or bleeding events. The selected medications included common ones like antihypertensives, antidiabetics, betablockers, statins, therapies with known association with anticoagulation such as NSAID, aspirin and CP450 inhibitors. This covariate information was combined into risk scores related to the outcomes under consideration, namely: (1) Charlson Index [3]) for death, (2) CHAD2DS2-Vasc score [16, 9] for stroke, and (3) HAS-BLED score [17] for bleeding. Other variables under consideration included patient age, sex, deprivation index as a proxy for lifestyle influence, type of AF, and if AF was first diagnosed in hospital vs. primary care. The complete covariate list and associated codes can be found in Supplementary Table 4.

#### Design

We followed a ‘per-protocol analysis’. Start date is the first date of oral anticoagulant prescription at or after AF diagnosis. If the date of first prescription took place within 90 days before the first diagnosis of AF, it was still considered to be related to this diagnosis. Patients exposed to oral anticoagulants within one year before this start date were excluded from the cohort. Patients with any outcome under consideration taking place within 15 days from the start date of follow-up were not included in the analyses since in an observational setting these endpoints cannot be attributed to treatment effects. A patient was considered lost to follow-up if (1) dies, (2) leaves the practice or the practice leaves CPRD or (3) at the end of the study period or (4) discontinue ‘assigned’ naive treatment; whichever happens first.

The follow-up time was divided into 3-month periods. At each time period: (a) baseline covariates were estimated from the 2-year medical history; (b) treatment *A* was set to *1* (or *0*) if at least one NOAC (or VKA) prescription was found during the time period; (c) censor was set to *1* if the patient was lost to follow up during the time period; (d) selected outcomes were set to *1* if they first took place during the time period and before censoring. These values where input into the causal inference algorithm.

### 2.3 Estimation of treatment effects

We aimed to measure the effectiveness of NOAC vs. VKA therapies for the prevention of stroke in AF patients as a difference in time-to-event probabilities, as well as to investigate the presence of heterogeneity of treatment effect. Following from previously validated methodology [6] we performed four steps:

Step 1 Initial estimation of treatment effect conditioned in each covariate vector using survival outcome models: Follow-up time is divided into 90-day intervals, outcome and censor data are expressed as binary counts over these intervals, and survival probabilities at each time for each covariate vector are estimated using a super learner framework in which a cross-validated ensemble of prediction algorithms is used to find an optimal fit for each treatment group. The treatment effect is defined as the difference in the estimated probabilities between NOAC and VKA patients at each time interval and for each covariate vector. This step was performed using the SuperLearner R package [18] with cross-validated algorithms “SL.earth”, “SL.glmnet”, and two different parametrizations of “SL.xgboost”. Follow-up time was capped 2 years, since the median follow up was around 21 months for all outcomes of interest.

Step 2 Estimation of Average Treatment Effect (ATE) using one-step Targeted Maximum Likelihood Estimation (TMLE): The initial survival model (Step 1) is updated to adjust for measured confounding using the propensity score (also estimated using a super learner framework). This step was performed using the MOSS R package [2].

Step 3: Initial identification of variables with potential heterogeneous treatment effect: We identify important variables of treatment effect heterogeneity for each outcome using a permutation-based variable selection approach in Bayesian Additive Regression Trees (BART) [1]. This step was performed using the BartMachine R package [7].

Step 4: When relevant, estimation of Conditional Average Treatment Effect (CATE) using one-step Targeted Maximum Likelihood Estimation (TMLE) over selected potentially heterogeneous subgroups as identified in Step 3. CATE is estimated over groups of patients defined by the important variables from Step 3. This step was performed following the same methodology as in Step 2.

## 3 Results

There were 20,270 total eligible patients: 13,159 (65%) new users of VKA and 7,111 (35%) new users of NOAC. The flowchart indicating the extraction of this study cohort as well as crude person-year rates for selected outcomes is displayed in Figure 1. All outcomes under consideration were relatively rare. The most common outcome was death with a person-year rate of 6% followed by bleeding (3%) and ischaemic event (2%). The median follow-up time was 21 months, therefore, to avoid bias associated with large censoring, the treatment effect computations were carried out only up to 2 years of follow-up.

**Figure 1:**
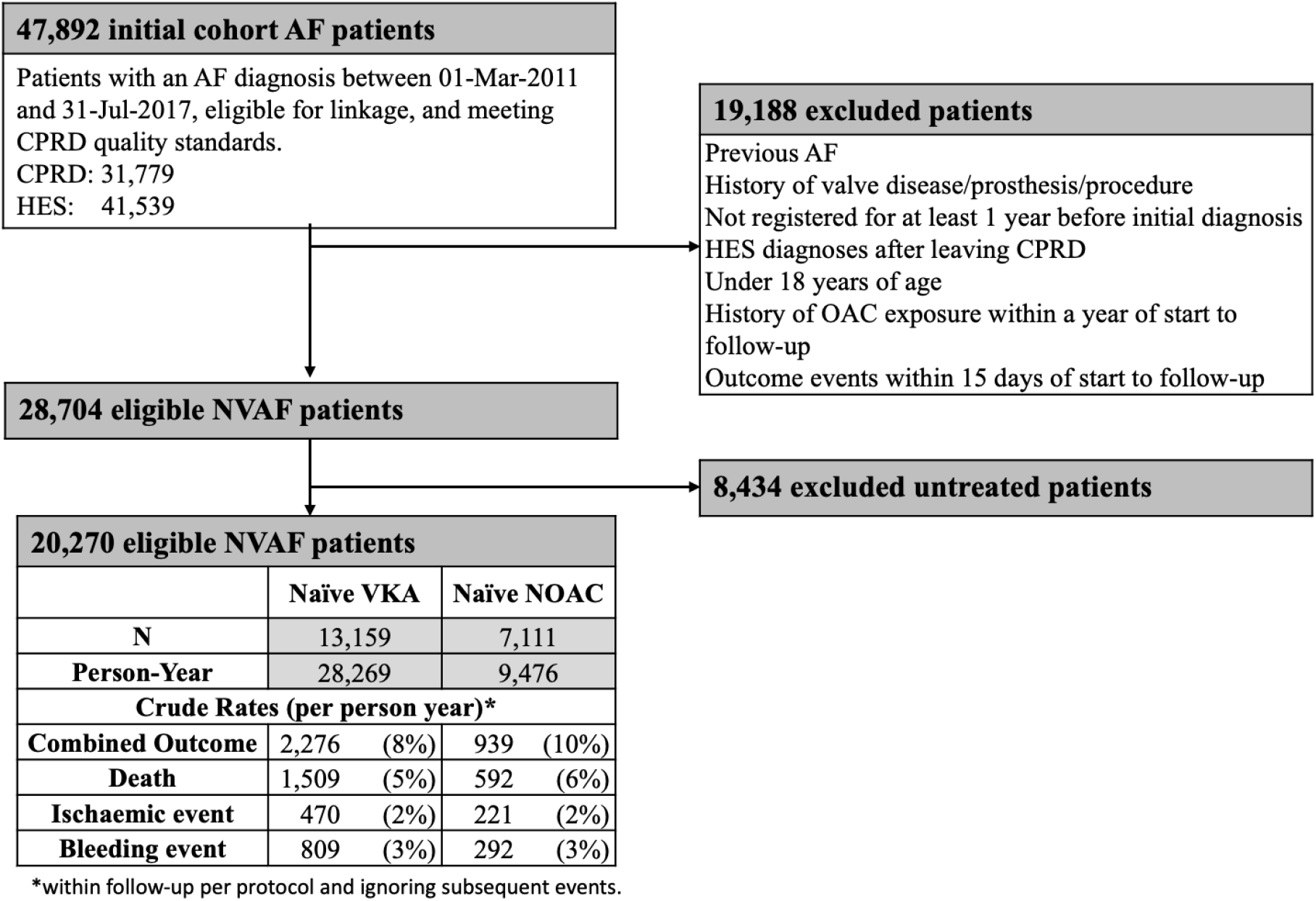
Cohort selection flowchart

### 3.1 Treatment assignment

Baseline characteristics at the start of follow-up are displayed in Table 1. Older patients, Paroxysmal AF, and an initial diagnosis of AF in acute care, were more prevalent in the NOAC group, as were several comorbidities such as hemiplegia, dementia, cerebrovascular disease, history of stroke and hypertension. The exceptions were Labile INR, end-stage renal failure, myocardial infarction and vascular disease, which were more common in the VKA treatment group. Use of VKAs as compared to NOACs was also found to be more common in patients with higher deprivation index. Baseline risk scores for death, bleeding and stroke had similar distributions across treatment groups, although patients in the NOAC group had on average a slightly larger Charlson and Chad2vasc2 scores and a slightly smaller Hasbled score. Antiplatelets, protom pump inhibitors and betablockers were more common amongst the NOAC group. This group also received medication on average later from first AF diagnosis. Figure 2 illustrates the overlap between VKA and NOAC patients as defined by their estimated propensity score at the start of follow-up. This treatment model had a good predictive performance, with a calibration slope of 1.06 (SD 0.07) and a c-index of 0.71 (SD 0.01) estimated via 5-fold cross validation.

**Figure 2:**
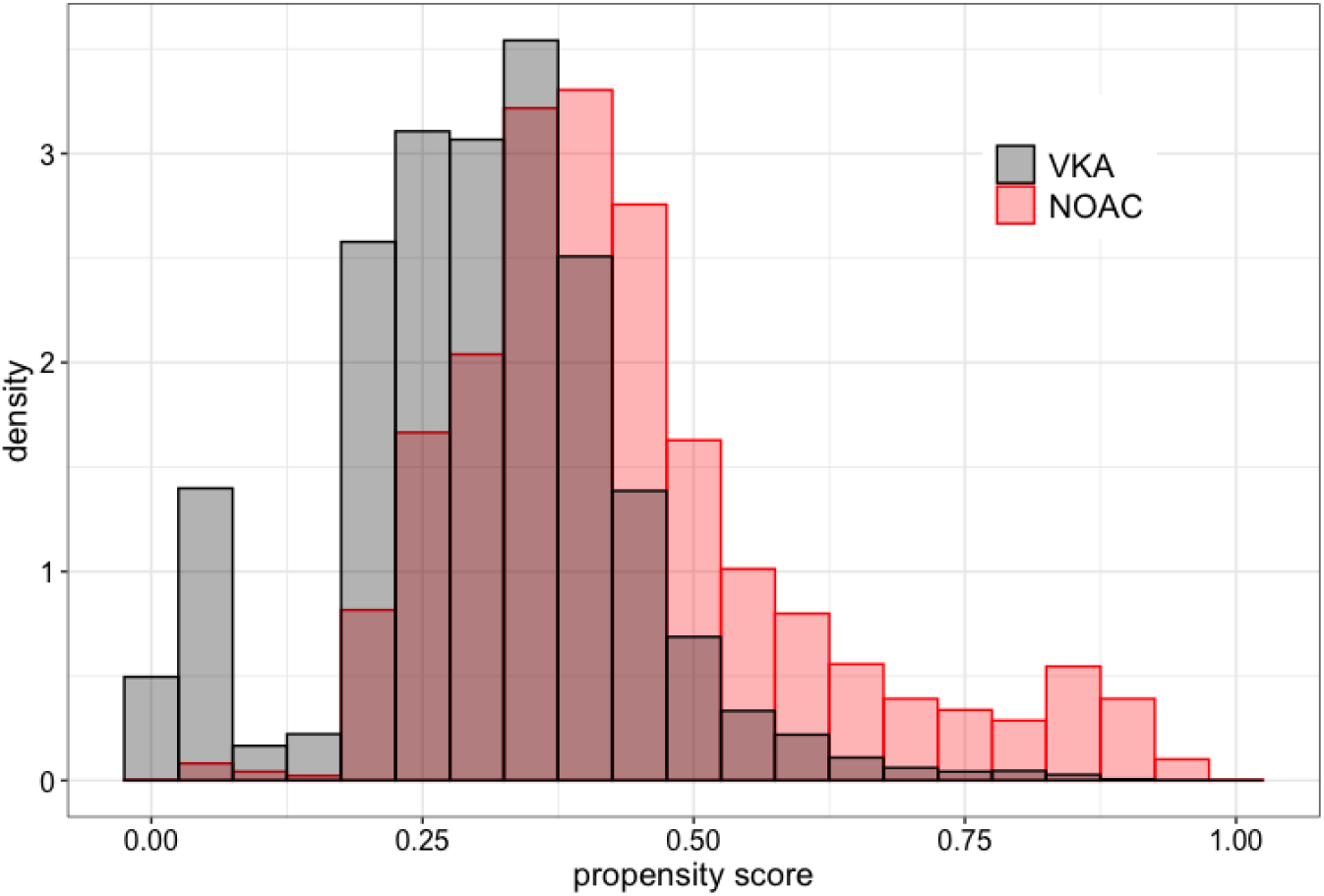
Propensity score at the start of follow-up

### 3.2 Treatment effects

Figure 3 shows the average treatment effects, *ATE* for the selected outcomes. The comparative effectiveness of NOAC vs VKA on all outcomes of interest, ischaemic event, major bleeding and death was not statistically or clinically significant. Averaged over the two years of follow-up, ATE was 0.00[95%0.00, 0.00] for ischaemic event, 0.00%[95% −0.01, 0.01] for major bleeding and 0.00[95% −0.01, 0.01] for death. The ATE was not sensitive to a reduction of the cohort to patients with a propensity score between 0.15 and 0.85, indicating that TMLE had adjusted for confounding as expected. The predictive performance of the outcome models was estimated using 5-fold cross validation. Concordance indices averaged between 6 months and 2 years were 0.76 (SD 0.04) for ischaemic event, 0.68 (SD 0.03) for major bleeding, and 0.75 (SD 0.02) for death.

**Figure 3:**
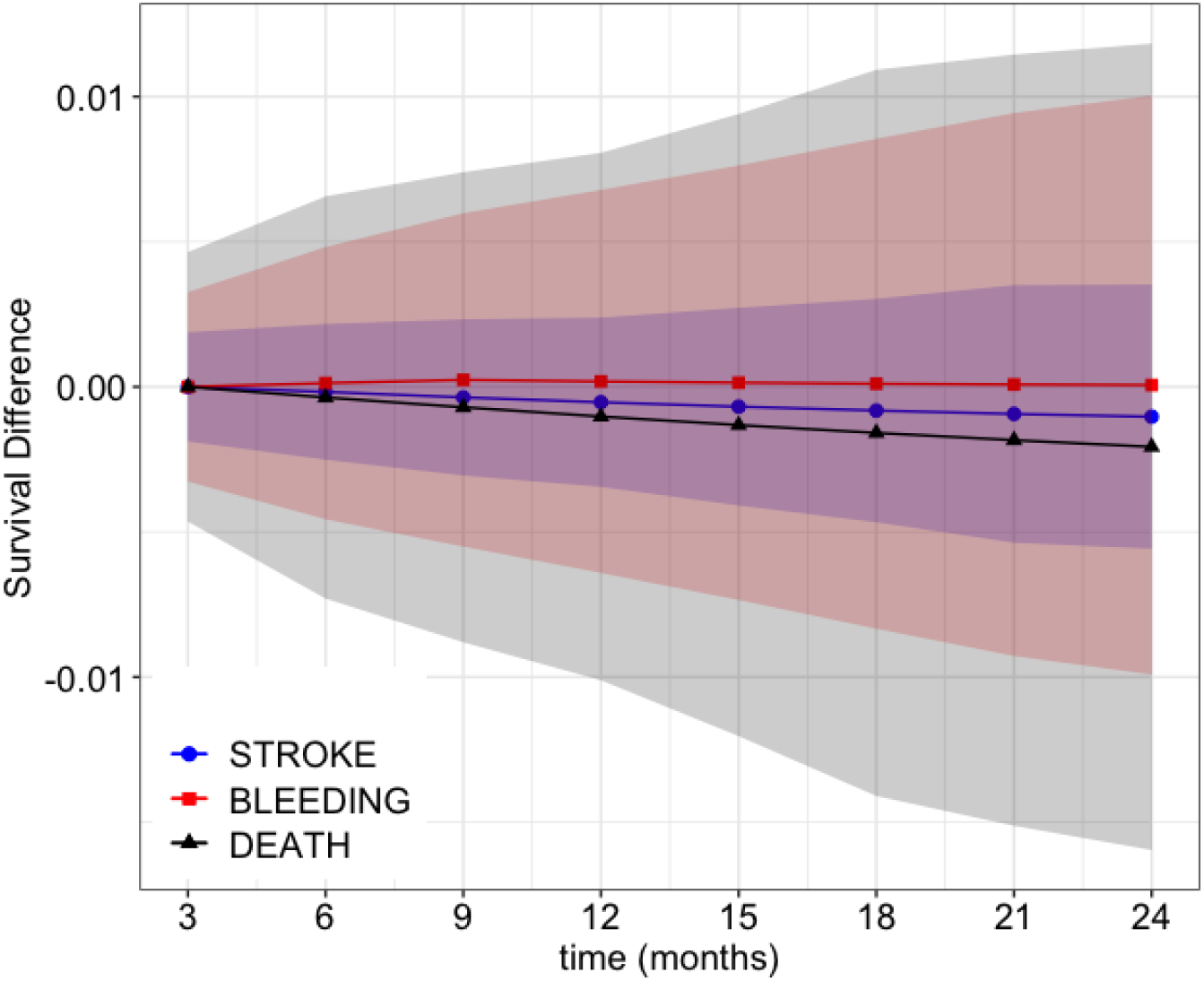
Estimated average treatment effect (ATE) for selected outcomes using TMLE adjusted potential outcomes from a SuperLearner model. The solid line corresponds to the average and the shaded areas correspond to the 95% confidence intervals.

The distribution of ITE for the selected outcomes is represented in Figure 4, which shows the estimated individual treatment effects averaged over follow-up time for patients with a baseline propensity score between 0.15 and 0.85. These distributions indicate a very small heterogeneity in treatment effect. When regressed on a BART model, a consistently important variable associated with a higher time-averaged ITE for major bleeding was history of major bleeding. However, the difference in associated CATE was really small (0.1% on average) and associated with high uncertainty and therefore did not warrant any further investigation. No covariates were found to influence the ITE distribution for ischaemic events or death.

**Figure 4:**
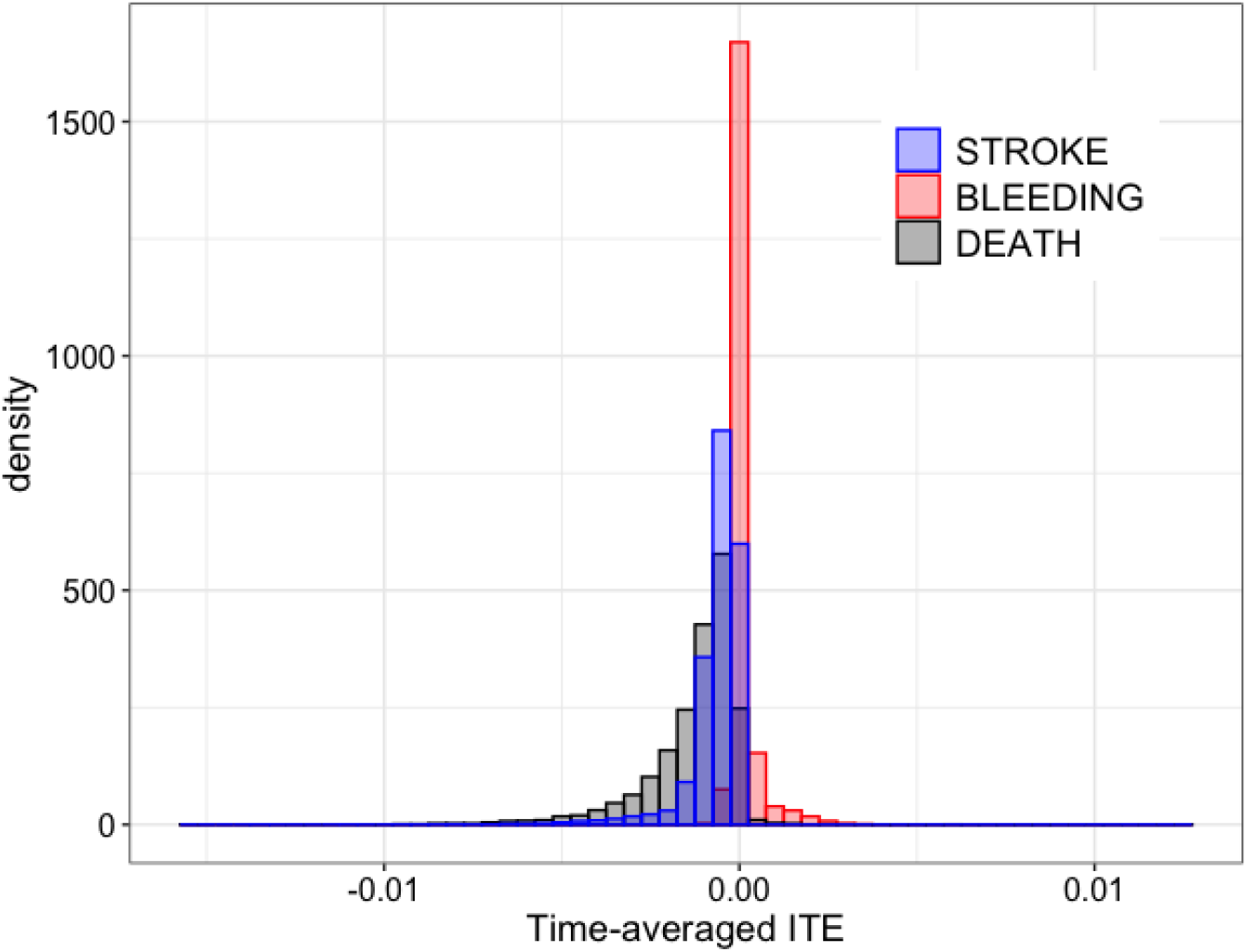
Distribution of time-averaged individual treatment effects (ITE) for selected outcomes using a SuperLearner model

## 4 Discussion

This study investigates the comparative effectiveness of NOACs vs VKAs for the prevention of stroke for patients with newly diagnosed non-valvular atrial fibrillation. In contrast with previous studies, treatment effect is defined using an absolute measure of risk as a function of medication exposure, namely the difference in time-to-event (survival) probability between NOAC and VKA treatment groups.

There was a significant patient similarity overlap between treatment groups although we observed some selective prescribing. NOAC users were more likely to be initially diagnosed in acute care, and were, in general, older, more complex patients. The exception were patients with end-stage renal failure, labile INR and history of myocardial infarction or vascular disease, who were more common in the VKA treatment group. VKA was also found to be more common in patients with higher deprivation index.

We found no statistically significant difference between NOACs and VKAs across the selected outcomes. This result generally agrees with the main findings from randomised clinical trials (see systematic review [19]) as well as with findings from previous studies in real-world settings (see systematic review [13] and posterior studies [10], [21], [5], [20], [4]), although in previous studies NOAC use (collectively) has been generally associated with similar or slightly lower risk of major bleeding.

In particular, four of the recent observational studies mentioned above also utilised the CPRD database (at least as one of their data inputs) for the comparative analysis of NOACs versus VKAs ([10], [21], [5], [20]). One study, Souverein et al 2020 [20], found that NOACs were associated with a slight increase in major bleeding events as well as in the risk of stroke (which in their case included haemorraghic stroke). They compared their results to those from Vinogradova et al 2018 [21] and found that the difference in the adjusted hazard ratios for major bleeding came from the risk associated with apixaban. This higher risk of major bleeding associated with apixaban was found in the CPRD dataset but not e.g. in the Danish dataset [20], and it was also not found in Van Ganse et al 2020 [4], where apixaban was associated with lower risk of major bleeding, stroke and mortality compared to VKAs.

No meaningful heterogeneity of treatment effect was found when looking at the distribution of the estimated ITE. This finding agrees with other secondary analyses of treatment effect heterogeneity from clinical trials [19], [12], but not with Pancholy et al [14], who found NOAC to be superior for women (but not men). Small treatment effect heterogeneities associated with patient’s sex [5], age [15], and type 2 diabetes status [23] have been reported in observational studies. However, these effects tended to be small and not confirmed by other studies.

## 5 Limitations

This study is limited by the nature and size of the data. In particular, exposure was determined from prescriptions by general practitioners missing direct information on treatment adherence as well as prescriptions from specialist clinics. Furthermore, no information on daily doses was contained in the version of the data used in this study. For these reasons, a simple, generous three-monthly time interval between prescriptions was utilised as a representation of medication adherence. Although this study makes use of a doubly-robust method for minimising the effect of confounders, residual confounding may still exist in the presence of unmeasured confounders. Individualised treatment effects were associated with large uncertainty and further analyses would require a larger dataset than the one used in this study.

## 6 Conclusions

No difference in effectiveness was found between naïve NOAC and naïve VKA users starting therapy upon an initial diagnosis of non-valvular atrial fibrillation. Analysis of heterogeneity of treatment effects for either rare diseases or common diseases with rare efficacy and/or safety outcomes require very large routinely collected datasets.

## Data Availability

Data are available on request from the CPRD. Their provision requires the purchase of a license, and our license does not permit us to make them publicly available to all. We used data from the version collected in February 2017 and have clearly specified the data selected in our Methods section.

## 7 Funding and ethics

This work was supported by National Health and Medical Research Council, project Grant No. 1125414. Ethics approval to use UK Clinical Practice Research Datalink data for this project was obtained from the Independent Scientific Advisory Committee (ISAC) of the Medicines and Healthcare products Regulatory Agency (MHRA), protocol number 17 093.

## Notes

### Competing Interest Statement

The authors have declared no competing interest.

### Author Declarations

Ethics approval to use UK Clinical Practice Research Datalink data for this project was obtained from the Independent Scientific Advisory Committee (ISAC) of the Medicines and Healthcare products Regulatory Agency (MHRA), protocol number 17 093.

